# Viral mutation, contact rates and testing: a DCM study of fluctuations

**DOI:** 10.1101/2021.01.10.21249520

**Authors:** Karl J. Friston, Anthony Costello, Guillaume Flandin, Adeel Razi

## Abstract

This report considers three mechanisms that might underlie the course of the secondary peak of coronavirus infections in the United Kingdom. It considers: (i) fluctuations in transmission strength; (ii) seasonal fluctuations in contact rates and (iii) fluctuations in testing. Using dynamic causal modelling, we evaluated the contribution of all combinations of these three mechanisms using Bayesian model comparison. We found overwhelming evidence for the combination of all mechanisms, when explaining 16 types of data. Quantitatively, there was clear evidence for an increase in transmission strength of 57% over the past months (e.g., due to viral mutation), in the context of increased contact rates (e.g., rebound from national lockdowns) and increased test rates (e.g., due to the inclusion of lateral flow tests). Models with fluctuating transmission strength outperformed models with fluctuating contact rates. However, the best model included all three mechanisms suggesting that the resurgence during the second peak can be explained by an increase in **effective contact rate** that is the product of a rebound of **contact rates** following a national lockdown and increased **transmission risk** due to viral mutation.

## Introduction

The secondary wave of SARS-CoV-2 prevalence in the United Kingdom is unfolding with a fluctuating time course that was not anticipated by epidemiological modelling. This begs the question, has something changed? Davies et al (Davies et al., 2020) present compelling evidence that a novel SARS-CoV-2 variant—Variant of Concern 202012/01 (a.k.a., lineage B.1.1.7)—is more transmissible than existing SARS-CoV-2 viruses in circulation. Combining multiple data sources and quantitative modelling they compared four competing biological mechanisms or hypotheses for why the new variant might be spreading more quickly. This report uses a similar approach to ask a broader question: to what extent does increased viral transmission explain the current resurgence during the secondary peak in the United Kingdom. We address this question by comparing the evidence for different hypotheses about how the available data, at a national level, are best explained. These hypotheses or models entail one or more mechanisms. The three mechanisms considered were, in order of plausibility:

### Mechanism 1

*fluctuations in viral transmission strength*. This has received attention recently based upon phylodynamic studies and statistical analyses of regional and cohort prevalence trends; particularly in the south-east of England (Davies et al., 2020). In brief, the idea here is that mutations affecting the spike protein render the virus more transmissible; possibly gaining access to cohorts or transmission reservoirs, such as young children. On this view, the current resurgence of prevalence is caused by increased **transmission risk** due to a new viral strain or genetic variant. The biological mechanisms considered in (Davies et al., 2020) included increased infectiousness, immune escape, increased susceptibility among children, and a shorter generation time. An alternative hypothesis is that increased **transmission risk** is due to nonviral characteristics, such as the regional variations in population density, seasonal fluctuations in temperature and humidity and so on (Mecenas, Bastos, Vallinoto, & Normando, 2020; Wu et al., 2020). On this view, a region’s dominant strain may appear more transmissible than another region’s dominant strain due to founder effects (Ruan et al., 2020). More nuanced versions of this explanation go beyond a founder effect and consider the competition for hosts, when the reproduction ratio of competing strains is near an unstable fixed point; namely, just above one (Anthony Brookes personal communication); see for example (Dimas Martins & Gjini, 2020; Nurtay, Hennessy, Sardanyés, Alsedà, & Elena, 2019). Nonviral characteristics bring us to the second mechanism.

### Mechanism 2

*seasonal fluctuations in contact rates.* This hypothesis focuses on the behaviour of the host—as opposed to the virus—in nuancing viral transmission. Technically, the **effective contact** rate is the product of the **total contact rate** and **transmission risk** or strength. This mechanism rests on the former. For example, seasonal or annual fluctuations in interpersonal contact (e.g., festive and religious events) or between communities (e.g., summer holidays, return to universities, school terms *et cetera*) may have a material effect on the rate at which the virus replicates. Under this hypothesis, it is changes in behaviour—and not the propensity of the virus to transmit itself—that affords the best explanation for the fluctuations in prevalence. Notice that this mechanism may operate independently of changes in transmission risk: for example, “we did not find evidence of differences in social interactions between regions of high and low VOC 202012/01 prevalence, as measured by Google mobility and social contact survey data.” (Davies et al., 2020). An alternative hypothesis is that neither contact rates nor transmission risks are changing and that the current resurgence is explained by changes in **testing** and **case definition**.

### Mechanism 3

*fluctuations in testing and reporting*. This mechanism can easily explain the recent increase in reported cases in terms of the increased testing rates—in particular, the inclusion of lateral flow device test results into the Pillar Two reported cases in the United Kingdom. These have only appeared in the past few weeks and now account for a substantial proportion of daily reported new cases (now—on 28 December 2020—at about 36,000 per day). Furthermore, the differential sensitivity and deployment of lateral flow and PCR tests may cause a change in positivity rates, in the absence of any change in prevalence. Although this is a plausible hypothesis for reported new cases, does it explain fluctuations in hospital admissions or ensuing fatalities? Perhaps counterintuitively, it can. This hypothesis rests upon the fact that the 28-day criteria—that defines a COVID-related death—depends upon the testing rate. This follows because there is a fluctuating probability of having a test by chance in the previous 28 days, which is conditionally independent of the probability of being infected with coronavirus. Clear evidence for this is the dissociation between the daily death rates based upon the 28-day testing criterion and deaths that are certified as being due to COVID-19. For example, in the United Kingdom, during the first wave, 1,356 people were dying every day at its peak due to COVID-19, whereas the 28-day peak death rate was 1,073 (i.e., 79% of certified deaths). *The situation reverses during the second peak* with daily rates of 482 and 505 respectively (i.e., 104% of certified deaths). In short, the 28-day case definition underestimated mortality rates in the first wave and overestimated them in the second (e.g., due to incidental PCR positive test results from patients with non-COVID-19 related admissions). The most parsimonious explanation for this is the increase in the probability of being tested in the past 28 days, irrespective of whether patients have COVID-19 or not^1^.

Each of the above mechanisms may operate in combination, leading to eight hypotheses or models, with and without each of the three mechanisms. To evaluate the evidence for these eight hypotheses, we used Bayesian model comparison and dynamic causal modelling.

### Dynamic causal modelling

The dynamic causal model used here is formally the same as described recently (K. J. Friston, Flandin, & Razi, 2020). However, in this DCM we included fast fluctuations in three key variables that model the above mechanisms. Dynamic causal modelling is particularly apt to address these kinds of questions. First, it an expressive (i.e., relatively assumption free) epidemiological model that supports formal model comparison (i.e., it does not rely upon cross validation or information criteria like the AIC, DIC or BIC). Second, in principle, it can resolve questions that involve circular causality: for example, does the emergence of a new viral strain cause an increase in viral transmission or does an increase in viral transmission cause the emergence of a new strain—or both? In principle, questions of this sort can only be resolved using a generative model with cyclic dependencies. Finally, dynamic causal modelling is routinely applied to fluctuating states in the form of stochastic DCM (Li et al., 2011).

The application dynamic causal modelling to the current outbreak has, to date, used deterministic states, namely the sufficient statistics of probability distributions over epidemiological states. However, there is another class of models where both the parameters and states are random variables, calling for inference about both the latent states generating data and the model parameters controlling the evolution of those states. Broadly speaking, there are three approaches to these kinds of models. First, one can estimate time-dependent probability distributions over parameters and latent states using variational procedures, sometimes referred to as generalised (variational) filtering (K. Friston, Stephan, Li, & Daunizeau, 2010). Second, one can treat the parameters as slowly fluctuating states and use conventional Bayesian filtering, for example unscented filters (Schiff, So, Chang, Burke, & Sauer, 1996). Finally, one can introduce fluctuations by expanding certain parameters using temporal basis functions (Daunizeau, Stephan, & Friston, 2012). Here, we choose the latter^2^.

To equip the model with fluctuations, we modulated three key parameters using discrete cosine transform (DCT) sets over a period of one year^3^. In other words, to model fluctuations in any given rate constant, we multiplied it by a linear mixture of cosine functions of time. The three parameters in question were chosen to embody the mechanisms above, namely:

### Changes in transmission strength (DCT order 8)

Transmission strength is defined as the probability of becoming infected after a close (i.e., effective) contact with an infectious person. This stands in for increased **transmission risk** due to viral mutation or any other changes in transmission strength, e.g., temperature, ventilation, seasonal variations in immunological status, probability of coinfection *et cetera*).

### The probability of visiting high contact risk location on any given day (DCT order 16)

This stands in for fluctuations in **contact rates** determined by social distancing behaviour and restrictions on the places people tend to visit (e.g., retail or hospitality venues, schools *et cetera*). This parameter models the host-dependent contributions to variations in transmission that model fluctuations around a prevalence-dependent social distancing, e.g., last-chance partying before— and rebound after—lockdown.

### Probability of being tested (DCT order 16)

This stands in for fluctuations in **testing rates** around the slow increases in rates, modelled as mixtures of sigmoid functions of time for Pillar One, Pillar Two and lateral flow testing.

Note that these fluctuations are around parameters or rate constants that are already time-dependent in the DCM. In other words, they are fast fluctuations that modulate parameters that change slowly: transmission strength is assumed to fluctuate over the seasons, using a cosine function with a period of one year. The probability of visiting a high-risk location (e.g., leaving home) is a non-linear function of prevalence, which models societal and governmental responses to increasing infection rates. Finally, the test rates are modelled with a series of sigmoid functions that are fitted to reported testing capacity. Please see (K. J. Friston et al., 2020) for details.

Implicit in the introduction of these fluctuations is a separation of timescales into slow trajectories that unfold over several months and faster fluctuations over several weeks. There is a principled reason for this separation which, in physics, is known as an adiabatic approximation (Koide, 2017). This inherits from the circular causality alluded to above, also known as the slaving principle (Haken, 1983). In brief, any dynamical system can be separated into slow (macroscopic) modes and fast (microscopic) modes. This separation rests upon an eigen-decomposition of the systems Jacobian. From our perspective, the interesting thing here is the circular causality induced by this decomposition, where slow modes are constituted by the average of microscopic variables, while the slow modes enslave the fast variables: c.f., the centre manifold theorem (Carr, 1981). In the present setting, this means that it is true to say that the fast emergence of a new strain subtends a peak of infection in a particular area while, at the same time, saying that the increasing prevalence enslaves—contextualises or creates the opportunity for—the emergence of a new strain^4^.

## Results

Bayesian model comparison—using a variational (free energy) approximation to log evidence— provided overwhelming evidence for the combination of all three mechanisms. This was the fourth model Figure 1, which shows the relative log evidence and probability distribution over the eight alternative models. In short, the evidence at hand suggests that we need all three mechanisms to explain the available data (listed in the legend to Figure 2). Interestingly, the next two most plausible models both involved fluctuations in transmission strength, with and without fluctuations in testing, speaking to a clear contribution of recent changes in transmission risk.

**Figure 1:**
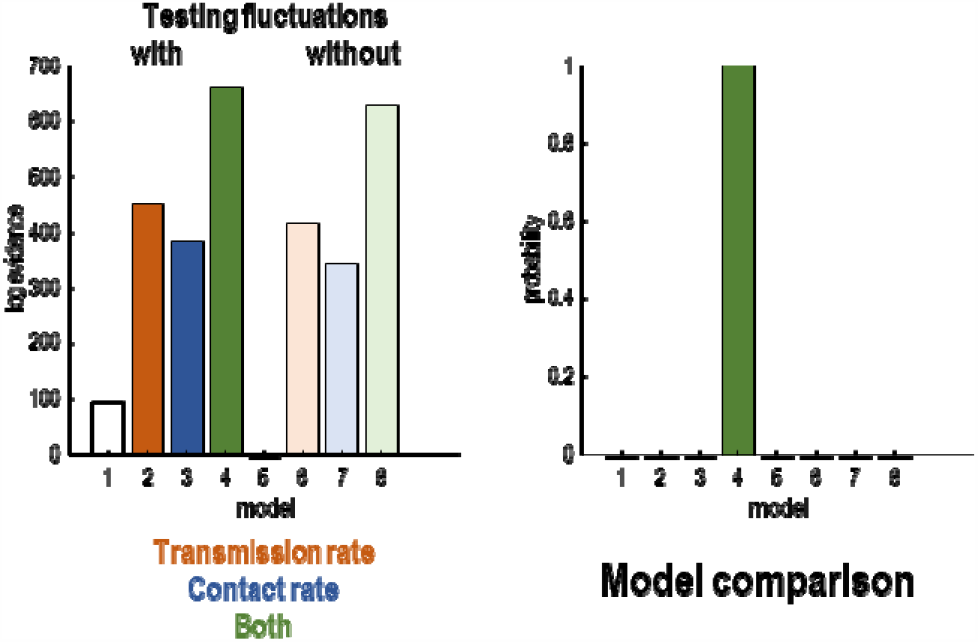
Bayesian model comparison of eight models created by allowing and disallowing three kinds of fluctuations: fluctuations in transmission rate, contact rate and testing rate. The left panel shows the log evidence for each model relative to the worst model (with no fluctuations). The right panel shows the equivalent results in terms of the probability density over models.

**Figure 2:**
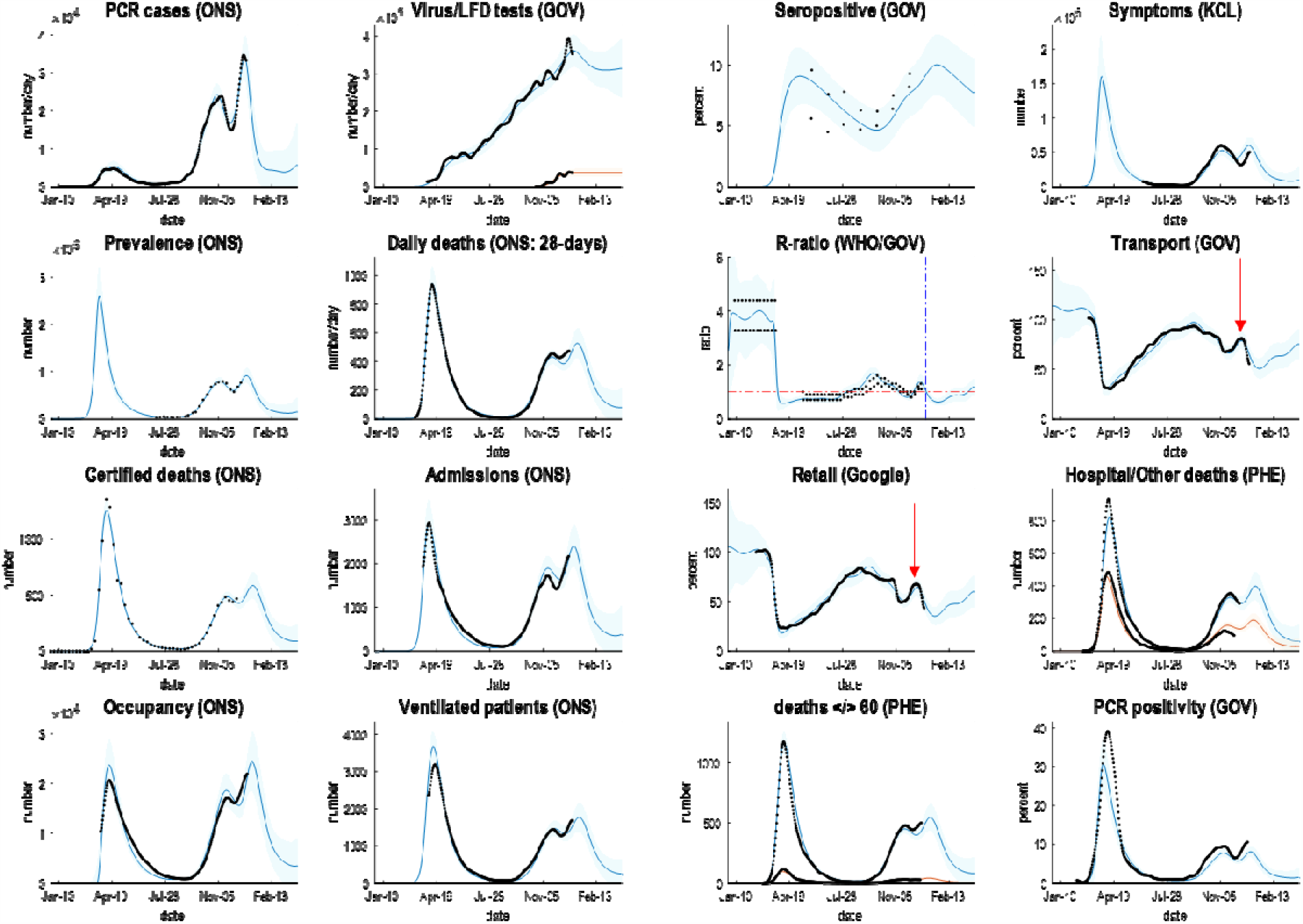
Posterior predictions of various outcomes, ranging from confirmed cases through to the reproduction ratio from the following sources: https://coronavirus.data.gov.uk; https://www.ons.gov.uk/peoplepopulationandcommunity/healthandsocialcare/conditionsanddiseases/datasets/coronaviruscovid19infectionsurveydata; https://covid.joinzoe.com/data#levels-over-time; https://www.gov.uk/guidance/the-r-number-in-the-uk#contents; https://www.gov.uk/government/statistics/transport-use-during-the-coronavirus-covid-19-pandemic; https://www.google.com/covid19/mobility/. The key thing to take from these results is the ability to provide a fairly accurate account of multiple aspects of the epidemic; ensuring an internal consistency to the explanations furnished by the model. The coloured lines correspond to posterior expectations, while the shaded areas correspond to 90% credible intervals. The black dots are the empirical timeseries used to estimate the model parameters and their posterior uncertainty.

The parameters of the full model—with all three mechanisms—are provided in Table 1, using the format described in (K. J. Friston et al., 2020). The interesting aspect of these results is summarised in Figures 2 and 3, which report posterior predictive densities over the outcomes used to fit the models and the underlying or latent epidemiological states. Because these models produce posterior predictive densities over all data space, including the future, they can be regarded as long-term forecasts (of missing data in the future). The dots correspond to the empirical timeseries.

**Table 1:** prior and posterior parameters

|  | Description | Prior mean | Lower bound | Upper bound | Posterior mean | Lower bound | Upper bound |
| --- | --- | --- | --- | --- | --- | --- | --- |
| 1 | population size ($M$) | 67.8 | 67.8 | 67.8 | **67.8** | 67.8 | 67.8 |
| 2 | initial cases | 0.006 | 0 | 1279 | **0.028** | 0.016 | 0.049 |
| 3 | pre-existing immunity | 0.1 | 0.083 | 0.12 | **0.10** | 0.08 | 0.12 |
| 4 | initially exposed | 0.1 | 0.08 | 0.12 | **0.10** | 0.08 | 0.13 |
| 5 | relative efflux | 0.1 | 0.054 | 0.183 | **0.077** | 0.042 | 0.142 |
| 6 | $P(\text{leaving home})$ | 0.36 | 0.33 | 0.39 | **0.36** | 0.34 | 0.39 |
| 7 | lockdown threshold | 0.01 | 0.005 | 0.018 | **0.011** | 0.010 | 0.012 |
| 8 | seropositive contribution | 0.2 | 0.10 | 0.36 | **0.18** | 0.16 | 0.20 |
| 9 | viral spreading (days) | 0.04 | 0.02 | 0.07 | **0.003** | 0.003 | 0.004 |
| 10 | admission rate (hospital) | 0.4 | 0.32 | 0.50 | **0.42** | 0.40 | 0.43 |
| 11 | admission rate (CCU) | 0.1 | 0.08 | 0.12 | **0.07** | 0.05 | 0.08 |
| 12 | distancing sensitivity | 3 | 2.4 | 3.7 | **2.1** | 1.8 | 2.4 |
| 13 | Effective contacts: home | 2 | 1.6 | 2.5 | **1.3** | 1.2 | 1.6 |
| 14 | Effective contacts: work | 20 | 16 | 24 | **24** | 21 | 28 |
| 15 | transmission (winter) | 0.3 | 0.24 | 0.37 | **0.28** | 0.24 | 0.31 |
| 16 | transmission (summer) | 0.3 | 0.24 | 0.37 | **0.22** | 0.19 | 0.25 |
| 17 | infected period (days) | 4 | 3.6 | 4.3 | **4.0** | 3.7 | 4.2 |
| 18 | infectious period (days) | 5 | 4.6 | 5.4 | **4.5** | 4.2 | 4.8 |
| 19 | loss of immunity (days) | 160 | 128 | 199 | **187** | 178 | 196 |
| 20 | resistance (late) | 0.2 | 0.18 | 0.21 | **0.22** | 0.20 | 0.24 |
| 21 | asymptomatic period (days) | 7 | 6.4 | 7.5 | **8.3** | 7.9 | 8.6 |
| 22 | symptomatic period (days) | 8 | 7.3 | 8.6 | **7.9** | 7.7 | 8.2 |
| 23 | ARDS period (days) | 6 | 5.5 | 6.5 | **6.1** | 5.8 | 6.3 |
| 24 | $P(\text{ARDS}\vert \text{symptoms}): \text{winter}$ | 0.01 | 0.009 | 0.010 | **0.0065** | 0.0062 | 0.0068 |
| 25 | $P(\text{ARDS}\vert \text{symptoms}): \text{summer}$ | 0.01 | 0.009 | 0.010 | **0.010** | 0.009 | 0.010 |
| 26 | $P(\text{fatality}\vert \text{ARDS}): \text{winter}$ | 0.5 | 0.46 | 0.54 | **0.58** | 0.55 | 0.61 |
| 27 | $P(\text{fatality}\vert \text{ARDS}): \text{summer}$ | 0.5 | 0.46 | 0.54 | **0.25** | 0.24 | 0.27 |
| 28 | *FTTI efficacy* | 0.036 | 0.033 | 0.039 | **0.036** | 0.033 | 0.039 |
| 29 | *testing bias (pillar one)* | 16 | 12.8 | 19.9 | **9.1** | 8.2 | 10.1 |
| 30 | *testing bias (pillar two)* | 8 | 6.4 | 9.9 | **5.2** | 4.7 | 5.7 |
| 31 | *test delay (days)* | 3 | 2.7 | 3.2 | **2.9** | 2.7 | 3.1 |
| 32 | *vaccination rate* | 0 | 0 | 0 | **0** | 0 | 0 |
| 33 | *false-negative rate* | 0.2 | 0.18 | 0.21 | **0.18** | 0.16 | 0.19 |
| 34 | *false-positive rate* | 0.002 | 0.001 | 0.002 | **0.002** | 0.0018 | 0.0022 |
| 35 | *testing: capacity* | 0.001 | 0.0005 | 0.0018 | **0.0014** | 0.0012 | 0.0016 |

Sources: https://royalsociety.org/-/media/policy/projects/set-c/set-covid-19-R-estimates.pdf and https://arxiv.org/abs/2006.01283:

**Figure 3:**
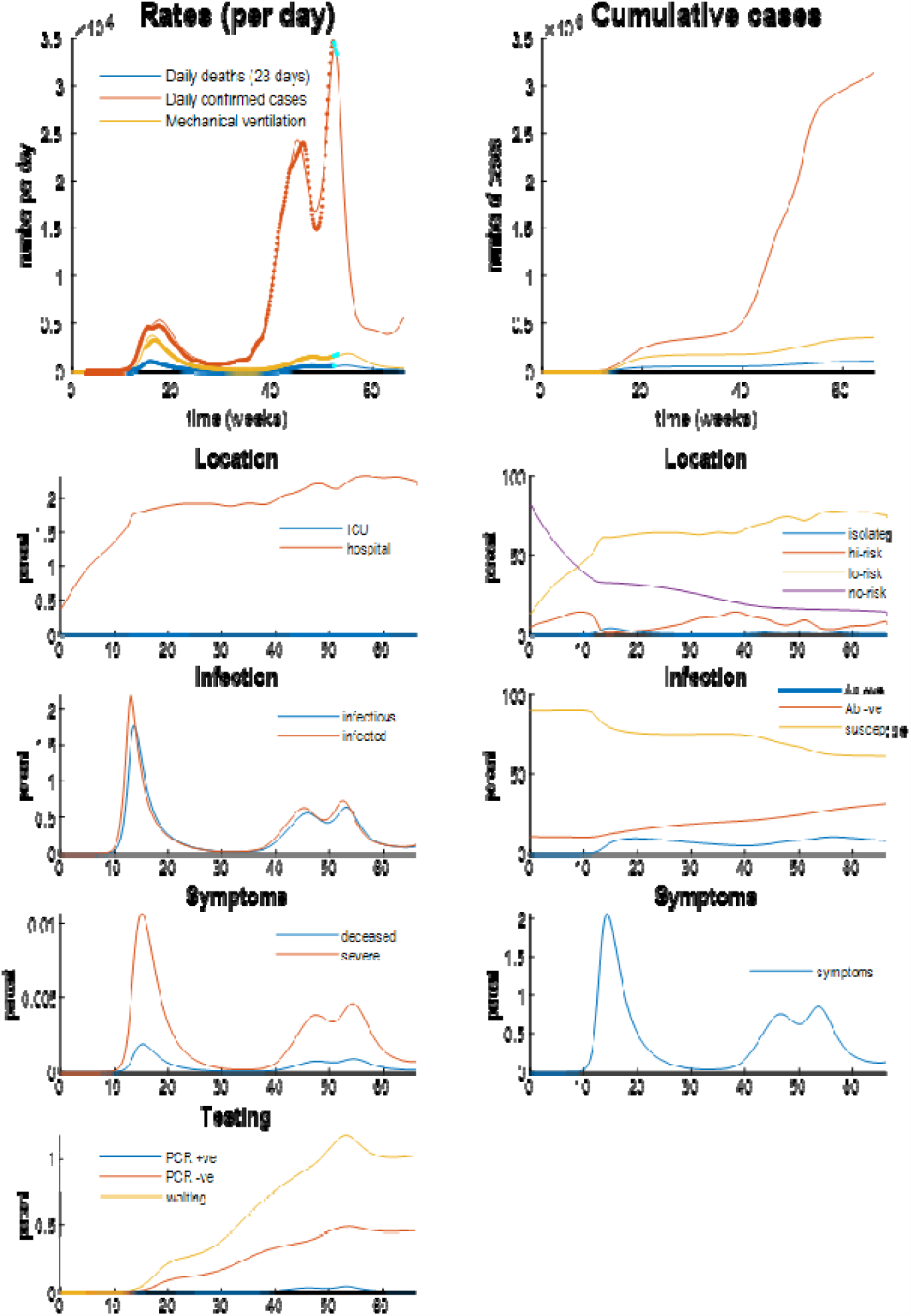
posterior predictions and underlying latent states. The upper panels show posterior expectations (coloured lines) and data (dots). The lower panels show the underlying posterior estimates of latent states generating these data. Please see (K. J. Friston et al., 2020) for a full explanation of the latent states and their interpretation.

It can be seen that this model provides an accurate account of many empirical aspects of the current outbreak; ranging from the number of new cases reported each day—through the official estimates of the reproduction ratio—to metrics of contact rates provided by the Department of Transport and Google estimates of mobility (here, ‘retail’ activity). Figure 4 shows the time varying transmission strength and contact risk. Recall from above, that these are slow changes in probabilities that are modulated by fast fluctuations, evincing peaks at various times over the past year.

**Figure 4:**
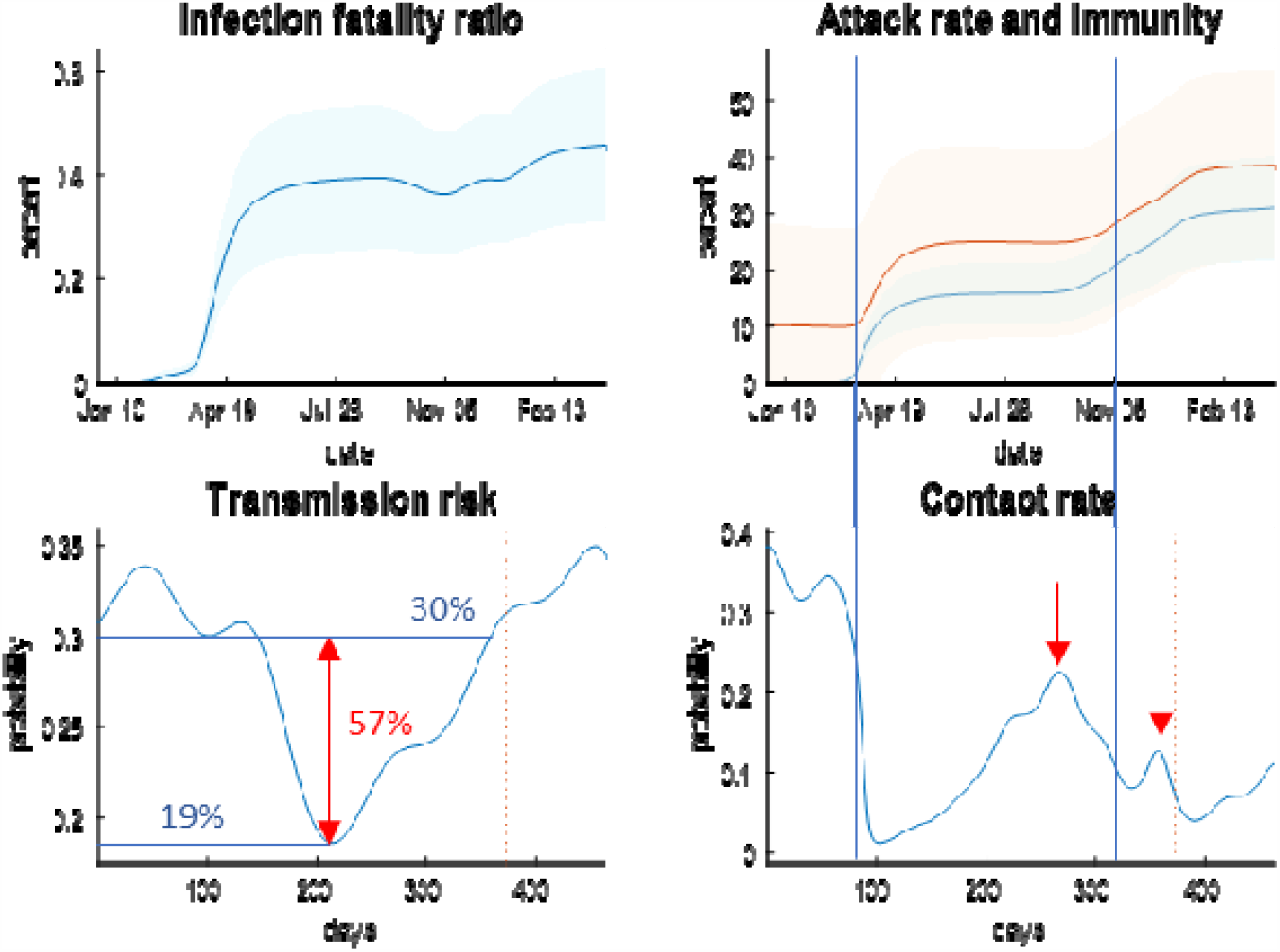
the **upper panels** report posterior predictions of cumulative infection fatality rates, attack rates and population immunity for the United Kingdom. The coloured lines correspond to posterior expectations, while the shaded areas correspond to 90% Bayesian credible intervals. The **lower panels** show the fluctuating time course of transmission strength and contact rate as parameterised by the probability of visiting a high-risk location each day. The vertical blue lines depict the onsets of the first and second national lockdowns in the United Kingdom.

Of particular interest is the recent peak in transmission rate that increased from about 20% in the summer to 30% in December: see Figure 4, lower left panel. This constitutes an increase of 57%, which is remarkably similar to the estimate of increased transmissibility of 56% reported in (Davies et al., 2020)^5^. This can be compared with a fluctuation in contact rates in the pre-Christmas period (lower right panel). This peak is less than the peak at the end of summer when students returned to university and children returned to school (and people returned from their summer holidays). These peaks are highlighted with red arrows in Figures 2 and3. The (onset of the) national lockdowns are shown with vertical lines, suggesting a rebound in contact rates after the second lockdown may have shaped the underlying epidemiology (see also below).

### Regional epilogue

The role of national lockdowns in shaping the trajectory of the second surge in the United Kingdom clearly rests on viral spread among communities and regions. An intuition about the contribution of fluctuations in contact rate and transmission risk—to region-specific trajectories—can be obtained by fitting the best model above to data from (lower tier) local authorities: in this instance, reported daily cases, deaths within 28 days of a positive PCR test and positivity rates as estimated by the Office of National Statistics, UK.

Figure 5 shows an example of two local authorities (Liverpool and Brentford) who experienced relatively early and late second peaks, respectively. These examples are based on region-specific parameter estimates (e.g., the number of initial cases, social distancing threshold, influx and efflux of people into the population, the typical number of people in a high and low-risk contact setting, et cetera). The remaining parameters (and fluctuations) were taken from the above analysis of the United Kingdom^6^, under the assumption they are conserved over local authorities (e.g., fluctuations in testing capacity, mobility related to holidays and schools, et cetera).

**Figure 5:**
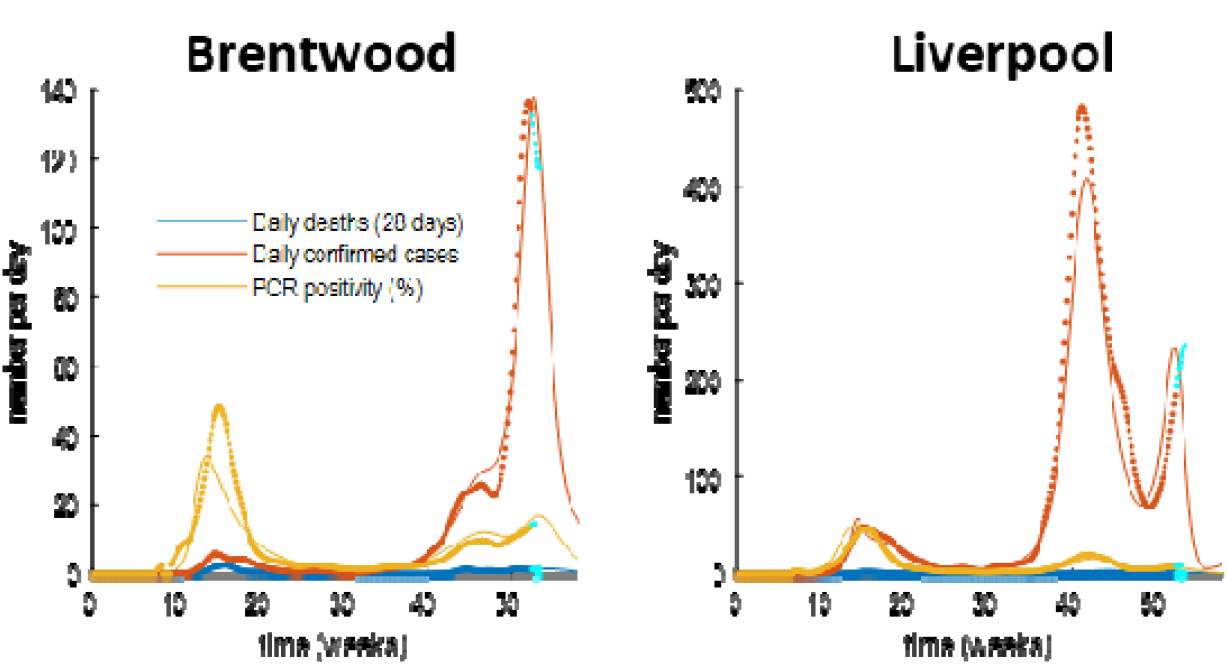
posterior predictions of daily deaths (within 28 days of a positive PCR test), daily confirmed cases reported, and positivity (percent), for two lower tier local authorities with late and early second peaks. Both show a double-hump profile but with distinct peaks that are separated by a month or so. The format of this figure follows the upper left panel in Figure 3. This analysis was repeated for all lower tier local authorities. The results are shown in 6.

Figure 6 shows the results of fitting the full model to 380 lower tier local authorities in the United Kingdom, and then presenting the ensuing timeseries as an image to illustrate the distribution of the timing of the second peak. The upper row shows the reported cases ranked separately for the first and second surges, while the second row shows the equivalent trajectories predicted by the model. The advantage of using predicted trajectories is that one can rollout into the future: here, by 64 days. The relative contribution of fluctuations in testing, contact rates and transmission risk—to the regional spread of the virus—can be assessed by successively removing these fluctuations. The effect of fluctuating testing rates can be removed by plotting the underlying (posterior) estimates of prevalence, with and without fluctuations in contact rate and transmission risk. The resulting distributions suggest that fluctuations in contact rates around the national lockdown are primarily responsible for dissecting the second surge into two peaks (compare the predicted prevalence with and without fluctuations in contact rates: highlighted in pink).

**Figure 6:**
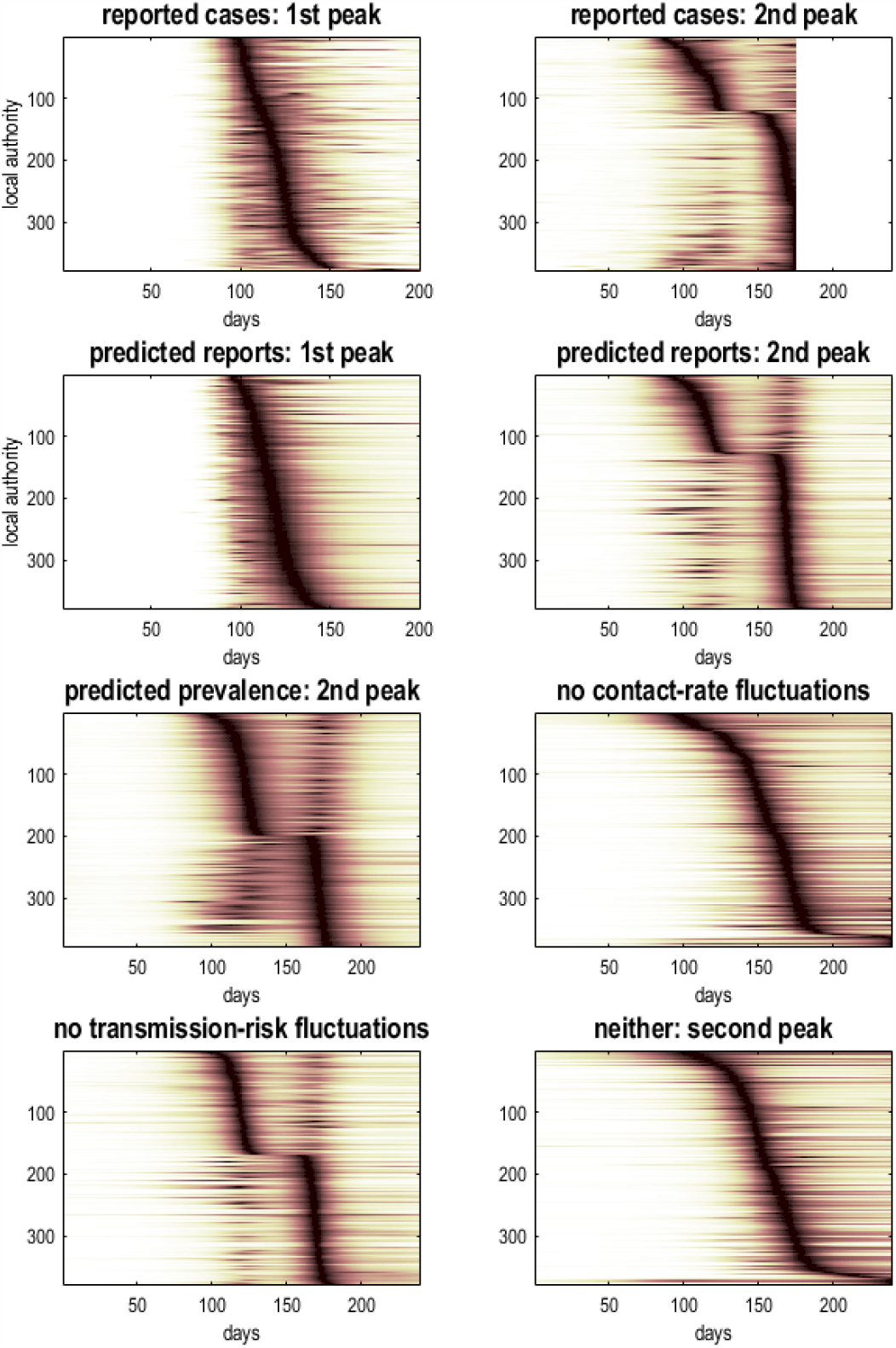
temporal raster or carpet plots based upon reported and predicted time courses of cases and prevalence for lower tier local authorities in the United Kingdom. The timeseries in each raster has been normalised to its peak value and then ranked according to the latency of the peak. This reveals the distribution of latencies over local authorities. The upper row is based upon reported confirmed virus tests, while the second row is based upon the corresponding posterior predictions, which include 64 days into the future. The lower panels report the underlying prevalence as estimated by the DCM that does not depend upon fluctuating testing rates. These predictions were evaluated with and without fluctuations in contact rates and transmission risk. The key result is the loss of a structured double peak when fluctuations in contact rates are removed, i.e., contact rates are driven just by the prevalence of infection, as opposed to any seasonal fluctuation (e.g., holidays, return to school et cetera).

About half the local authorities had a high amplitude initial surge that was suppressed by the national lockdown and followed by an attenuated resurgence. Conversely, regions with a smaller initial surge experienced a post-lockdown resurgence that exceeded their initial peak. Note that the predicted course of the trajectory into the future assumes a subsequent restriction of contact rates that is mediated by further lockdowns or mitigations.

## Conclusion

As might be expected, the explanations for fluctuations in reported cases and death rates witnessed recently in the United Kingdom rest upon the interaction among multiple factors. The model used here suggests that there are at least three factors in play. These include fluctuations in the transmission strength of the virus, fluctuations in exposure to the virus due to human behaviour and fluctuations in testing rates—that affect the way we measure mortality. The implications of this kind of analysis are straightforward, and recapitulate what most people have been recommending for the past year:

### Containment

namely, use every available resource—including surveillance testing—to identify regional hotspots so that the virus can be contained within a local area or outbreak cluster. This speaks to regional and context-sensitive responses (e.g., a tier system) with a special focus on restricting between-community mixing (e.g., using quarantine or travel restrictions). However, it does not necessarily speak to the indiscriminate use of national lockdowns, in the sense that the timing of a national lockdown may be optimal for one region but not another. For example, the first lockdown may have suppressed transmission in London but terminated prematurely for Manchester, where the prevalence of infection may not have been suppressed before unlocking. Conversely, a second lockdown may have been optimal for Manchester but too soon for London; in the sense that unlocking was premature and allowed the emergence of new strains of the virus in a region with rebounding contact rates. As noted by (Davies et al., 2020): “School closures in January 2021 may delay the peak and decrease the total burden in the short term. However, implementation of more stringent measures now with a subsequent lifting of these restrictions in February 2021 leads to a bigger rebound in cases, particularly in those regions that have been least affected so far”. It should be noted that these speculations do not consider the putative effects of vaccination, which will clearly affect contact rates by reducing the number of people who can transmit the virus.

### Suppression

this rests upon conventional public health measures and proven approaches to the control of infectious diseases. When specialised to the current situation, it speaks to a proper integration of NHS Test and Trace with local contact tracing teams.

### Elimination

this is now in sight, with the advent of effective vaccines.

There is clear consilience between the conclusions of the Bayesian model comparison above and the analyses reported in (Davies et al., 2020). The only real difference rests upon the way that mitigations are handled when predicting long-term outcomes. The dynamic causal model effectively combines an epidemiological model (formally not dissimilar to the one used in Davies et al., *ibid*) with an agent-based behavioural model to predict behavioural responses. In other words, it predicts behavioural (and governmental) responses based upon what has happened in the past— modelling the circular causality where viral transmission causes changes in behaviour (e.g., moving from one tier to another when prevalence exceeds a threshold) and where behaviour causes changes in viral transmission (e.g., precluding movement between different tiers). This means that the contact rates used in epidemiological models are replaced by the probability of encountering someone who is infected that depends upon behavioural responses.

Davies et al. (*ibid*) base their predictions on several scenarios involving lockdowns and school closures in January and February, with a focus on the rebound in fatalities, peaking in February through to April. The dynamic causal modelling predicts that a national lockdown (or extended tier 4 restrictions) will be implemented in the near future; although not as complete or extensive as the first national lockdown. The DCM further predicts that these restrictions will be relaxed slowly, after death rates start to fall in January. Crucially, these predictions rest on the assumption that transmission strength fluctuates and *can go down again*. This may or may not be a plausible assumption—and rests on the factors that underwrite changes in transmission (e.g., the viral characteristics considered by Davies et al., (ibid) and non-viral factors, such as the time spent outdoors, ventilation, *et cetera*).

This speaks to an important question, is the new strain here to stay or is it too transmissible for its own good?—as hinted at by considerations from evolutionary virology (Ignacio-Espinoza, Ahlgren, & Fuhrman, 2020; Peng et al., 2017; Schenk, Schulenburg, & Traulsen, 2020; Woods et al., 2011). In principle, this question can be addressed using model comparison, when the course of the epidemic reveals itself over the next month or two.

### Software note

The figures in this report can be reproduced using annotated (MATLAB) code available as part of the free and open-source academic software SPM (https://www.fil.ion.ucl.ac.uk/spm/), released under the terms of the GNU General Public License version 2 or later. The routines are called by a demonstration script that can be invoked by typing >> DEM_COVID_UK at the MATLAB prompt. At the time of writing (28-12-2020), these routines are undergoing software validation in our internal source version control system—that will be released in the next public release of SPM (and via GitHub at https://github.com/spm/). In the interim, please see https://www.fil.ion.ucl.ac.uk/spm/covid-19/.

## Data Availability

All the data used is from publically available database.

## Data availability

The data used in this technical report are available for academic research purposes from the sites listed in the legend to Figure 2.

## Acknowledgements

The Wellcome Centre for Human Neuroimaging is supported by core funding from Wellcome [203147/Z/16/Z]. A.R. is funded by the Australian Research Council (Refs: DE170100128 and DP200100757) and Australian National Health and Medical Research Council Investigator Grant (Ref: 1194910). We are especially grateful to Deenan Pillay for guidance on virology.

The authors declare no conflicts of interest.

## Footnotes

1 Technically, when one compares models with and without a test rate-dependent contribution to non-certified deaths and hospital admissions, there is overwhelming evidence for an effect of test rates (data not shown).

2 Strictly speaking, fluctuations are usually fast random effects that show no serial correlations (for example, the Wiener process in a Langevin equation). However here, we wanted smooth (analytic) fluctuations, where the smoothness can be controlled through the order or number of temporal basis functions.

3 A discrete cosine set is like a Fourier basis set and is the transformation of choice in signal processing and data compression. The number of basis functions, called the order, determines the highest frequencies and therefore the smoothness of fluctuations expressed as a mixture of cosine functions of time.

4 We have deliberately (over) used the word ‘fluctuation’ because the density dynamics in stochastic and statistical thermodynamics rest upon almost exclusively on fluctuation theorems (Evans & Searles, 2002; Seifert, 2012)

5 Note that these estimates refer to different things: Davies et al. (ibid) report a careful comparison of the transmissibility of different viral strains. The current quantification of transmissibility lumps together all SARS-CoV-2 strains and quantifies changes in transmission risk over time.

6 Technically, these estimates play the role of *empirical priors*; namely, posterior estimates from a prior analysis.

## Notes

### Competing Interest Statement

The authors have declared no competing interest.

## References

Carr, J. (1981). Applications of Centre Manifold Theory. Berlin: Springer-Verlag.

Daunizeau, J., Stephan, K. E., & Friston, K. J. (2012). Stochastic dynamic causal modelling of fMRI data: should we care about neural noise? Neuroimage, 62(1), 464–481. doi:10.1016/j.neuroimage.2012.04.061

Davies, N. G., Barnard, R. C., Jarvis, C. I., Kucharski, A. J., Munday, J. D., Pearson, C. A. B., … Edmunds, W. J. (2020). Estimated transmissibility and severity of novel SARS-CoV-2 Variant of Concern 202012/01 in England. medRxiv, 2020.2012.2024.20248822. doi:10.1101/2020.12.24.20248822

Dimas Martins, A., & Gjini, E. (2020). Modeling Competitive Mixtures With the Lotka-Volterra Framework for More Complex Fitness Assessment Between Strains. Frontiers in Microbiology, 11 (2132). doi:10.3389/fmicb.2020.572487

Evans, D. J., & Searles, D. J. (2002). The fluctuation theorem. Advances in Physics, 51(7), 1529–1585. doi:10.1080/00018730210155133

Friston, K., Stephan, K., Li, B., & Daunizeau, J. (2010). Generalised Filtering. Mathematical Problems in Engineering, vol., 2010, 621670.

Friston, K. J., Flandin, G., & Razi, A. (2020). Dynamic causal modelling of mitigated epidemiological outcomes. 2011.12400. Retrieved from https://ui.adsabs.harvard.edu/abs/2020arXiv201112400F

Haken, H. (1983). Synergetics: An introduction. Non-equilibrium phase transition and self-selforganisation in physics, chemistry and biology. Berlin: Springer Verlag.

Ignacio-Espinoza, J. C., Ahlgren, N. A., & Fuhrman, J. A. (2020). Long-term stability and Red Queen-like strain dynamics in marine viruses. Nature Microbiology, 5(2), 265–271. doi:10.1038/s41564-019-0628-x

Koide, T. (2017). Perturbative expansion of irreversible work in Fokker–Planck equation à la quantum mechanics. Journal of Physics A: Mathematical and Theoretical, 50(32), 325001. Retrieved from http://stacks.iop.org/1751-8121/50/i=32/a=325001

Li, B., Daunizeau, J, Stephan, E K., Penny, W., … Friston, K. (2011). Generalised filtering and stochastic DCM for fMRI. Neuroimage, 58(2), 442–457.

Mecenas, P., Bastos, R. T. d. R. M., Vallinoto, A. C. R., & Normando, D. (2020). Effects of temperature and humidity on the spread of COVID-19: A systematic review. PLoS ONE, 15(9), e0238339. doi:10.1371/journal.pone.0238339

Nurtay, A., Hennessy, M. G., Sardanyés, J., Alsedà, L., & Elena, S. F. (2019). Theoretical conditions for the coexistence of viral strains with differences in phenotypic traits: a bifurcation analysis. Royal Society open science, 6(1), 181179–181179. doi:10.1098/rsos.181179

Peng, G., Yang, Y., Pasquarella, J. R., Xu, L., Qian, Z., Holmes, K. V., & Li, F. (2017). Structural and Molecular Evidence Suggesting Coronavirus-driven Evolution of Mouse Receptor. The Journal of biological chemistry, 292(6), 2174–2181. doi:10.1074/jbc.M116.764266

Ruan, Y., Luo, Z., Tang, X., Li, G., Wen, H., He, X., … Wu, C.-I. (2020). On the founder effect in COVID-19 outbreaks – How many infected travelers may have started them all? National Science Review. doi:10.1093/nsr/nwaa246

Schenk, H., Schulenburg, H., & Traulsen, A. (2020). How long do Red Queen dynamics survive under genetic drift? A comparative analysis of evolutionary and eco-evolutionary models. BMC Evolutionary Biology, 20(1), 8. doi:10.1186/s12862-019-1562-5

Schiff, S. J., So, P., Chang, T., Burke, R. E., & Sauer, T. (1996). Detecting dynamical interdependence and generalized synchrony through mutual prediction in a neural ensemble. Phys Rev E Stat Phys Plasmas Fluids Relat Interdiscip Topics, 54(6), 6708–6724.

Seifert, U. (2012). Stochastic thermodynamics, fluctuation theorems and molecular machines. Rep Prog Phys, 75(12), 126001. doi:10.1088/0034-4885/75/12/126001

Woods, R. J., Barrick, J. E., Cooper, T. F., Shrestha, U., Kauth, M. R., & Lenski, R. E. (2011). Second-Order Selection for Evolvability in a Large Escherichia coli Population. Science, 331(6023), 1433–1436. doi:10.1126/science.1198914

Wu, Y., Jing, W., Liu, J., Ma, Q., Yuan, J., Wang, Y., … Liu, M. (2020). Effects of temperature and humidity on the daily new cases and new deaths of COVID-19 in 166 countries. Science of The Total Environment, 729, 139051. doi:https://doi.org/10.1016/j.scitotenv.2020.139051

